# Acceptability of the Venting Wisely Pathway for use in Critically ill Adults with Hypoxemic Respiratory Failure and Acute Respiratory Distress Syndrome (ARDS): A qualitative study protocol

**DOI:** 10.1101/2023.04.21.23288685

**Authors:** Karla Krewulak, Gwen Knight, Andrea Irwin, Jeanna Morrissey, Henry T. Stelfox, Sean Bagshaw, Danny J. Zuege, Amanda Roze des Ordons, Kirsten M. Fiest, Ken Kuljit S Parhar

## Abstract

**Introduction:** Hypoxemic respiratory failure (HRF) affects nearly 15% of critically ill adults admitted to an intensive care unit (ICU). An evidence based, stakeholder informed multidisciplinary care pathway (*Venting Wisely*) was created to standardize the diagnosis and management of patients with HRF and Acute Respiratory Distress Syndrome (ARDS). Successful adherence to the pathway requires a coordinated team-based approach by the clinician team. The overall aim of this study is to describe the acceptability of the *Venting Wisely* pathway among critical care clinicians. Specifically, this will allow us to: 1) better understand the user’s experience with the intervention and 2) determine if the intervention was delivered as intended.

**Methods and analysis:** This qualitative study will conduct focus groups with nurse practitioners, physicians, registered nurses, and registered respiratory therapists from 17 Alberta ICUs. We will use template analysis to describe the acceptability of a multi component care pathway according to seven constructs of acceptability: 1) Affective attitude; 2) Burden; 3) Ethicality; 4) Intervention coherence; 5) Opportunity costs; 6) Perceived effectiveness; and 7) Self-efficacy. This study will contribute to a better understanding of the acceptability of the *Venting Wisely* pathway. Identification of areas of poor acceptability will be used to refine the pathway and implementation strategies as ways to improve adherence to the pathway and promote its sustainability.

**Ethics and dissemination:** The study was approved by the University of Calgary Conjoint Health Research Ethics Board (CHREB). The results will be submitted for publication in a peer-reviewed journal and presented at a scientific conference.

**STRENGTHS AND LIMITATIONS:**

- This qualitative study will provide vital information about why the implementation of the *Venting Wisely* pathway may or may not have worked as anticipated.
- Findings will identify opportunities to improve pathway adherence and provide insights on how to sustain the intervention and scale to other sites.
- Acceptance and adherence of the *Venting Wisely* pathway has the potential to increase and standardize the use of evidence-informed, life-saving therapies for mechanically ventilated patients; this may improve outcomes and save costs to the healthcare system.
- Focus groups will be conducted with a wide variety of clinicians (nurse practitioners, physicians, registered nurses, registered respiratory therapists), and within various intensive care units (general systems, cardiovascular surgery, and neurosciences), and hospitals (regional, community, and tertiary).
- The study is being conducted in one province in Canada, which may limit generalizability.

## INTRODUCTION

Hypoxemic Respiratory Failure (HRF) is a common medical emergency affecting up to 15% of ICU admissions (1, 2). The most severe subtype of HRF is ARDS (3). ARDS is associated with significant mortality (over 30% in severe cases), functional disability, and increased health care resource utilization (4-10). Guideline-recommended approaches for the application of mechanical ventilation and adjunctive therapies for HRF and ARDS exist (11-15). Unfortunately, despite this, HRF and ARDS remain underdiagnosed and evidence-based interventions remain underutilised (10).

Effective clinical management of complex conditions such as HRF and ARDS requires a coordinated, multidisciplinary approach. The Institute of Medicine suggests using care pathways to coordinate and improve care of complex conditions (16). We developed a care pathway for HRF and ARDS called *Venting Wisely* that is evidence-informed, multidisciplinary and stakeholder derived (17). This pathway standardizes the diagnosis and management of HRF and ARDS. It includes 42 elements, however is focused on five key evidence-informed steps including measuring a patient’s height to estimate the size of their lungs, screening for hypoxemic respiratory failure daily, instituting lung protective ventilation consistently, and using neuromuscular blockade and prone positioning when indicated (17).

Acceptability of the intervention among clinicians is a crucial attribute for its’ success. Sekhon et al. define acceptability as a multi-faceted construct and propose the theoretical framework of acceptability (TFA) to evaluate the acceptability of healthcare interventions (18). The TFA consists of seven components: 1) *affective attitude* (how a clinician feels about the intervention), 2) *burden* (the clinician’s perception about the required amount of effort to participate in the intervention), 3) *ethicality* (the extent to which the intervention aligns with a clinician’s value system), 4) *intervention coherence* (the extent to which the clinician understands the intervention), 5) *opportunity costs* (benefits or costs to the clinician for using the pathway), 6) *perceived effectiveness* (the extent to which the clinician perceives the intervention as likely to achieve its purpose), and 7) *self-efficacy* (the clinician’s confidence that they can use the pathway).

Acceptability is recognized as an important implementation outcome that should be assessed in any complex intervention (19). Therefore, understanding the acceptability of the *Venting Wisely* pathway is important to understand the user’s experience of the intervention and whether the intervention is being provided as intended. Implementation of the *Venting Wisely* pathway is complex because it requires engagement of multidisciplinary intensive care unit (ICU) care team members, including nurse practitioners, physicians, registered nurses, or registered respiratory therapists. Our understanding of why the implementation of the *Venting Wisely* pathway does or does not work as anticipated will identify opportunities to improve pathway adherence and provide insights on how to sustain the intervention and scale to other sites (18). Strong acceptability of the *Venting Wisely* pathway has the potential to increase and standardize the use of evidence-informed, life-saving therapies for HRF and ARDS, improve patient outcomes, and reduce costs within the healthcare system. Study findings may also provide insights into how other complex interventions should and should not be implemented and adopted by multidisciplinary teams. The *Venting Wisely* pathway is currently being implemented through a cluster randomized stepped wedge trial (ClinicalTrials.gov NCT04744298) across 17 adult ICUs in Alberta, Canada as part of a hybrid implementation-effectiveness trial (20, 21).

## Objective

The overall objective of this study is to explore clinician perceptions of the **acceptability** of the *Venting Wisely* pathway among ICU clinicians in a diversity of ICUs. These data will inform iterative refinements of the pathway and the implementation strategy for this pathway and suggestions for facilitating pathway fidelity, sustainability, and scalability.

## METHODS AND ANALYSIS

### Study design

The study will be reported according to the Consolidated criteria for Reporting Qualitative research (COREQ)(22). The study was designed with input from a patient partner. Following implementation of the *Venting Wisely* pathway, we will conduct focus groups with critical care clinicians (nurse practitioners, physicians, registered nurses, or registered respiratory therapists involved in using the pathway). The full study protocol has been posted publicly (20). This focus group protocol has been posted on a pre-print server (https://www.medrxiv.org/) prior to completion of recruitment. Focus groups were initiated in April 2022. The target for study completion is late 2023.

### Participants and sampling frame

Participants will be eligible if they are a clinician (nurse practitioner, physician, registered nurse, or registered respiratory therapist) working in one of the 17 ICUs in Alberta, Canada that has experienced the implementation of the *Venting Wisely* pathway for at least two months. Inclusion criteria includes ICUs demonstrating adherence to key pathway elements (i.e., composite fidelity score of >70% or 10% gain above baseline) to ensure that focus group participants have adequate exposure to the pathway. Any ICU that does not meet these criteria will have their focus group conducted at the end of the study.

Eligible clinicians will be emailed a letter explaining the purpose of the study by the ICU manager or site pathway champions (*online supplemental file 1)*. Interested clinicians will be invited to contact the research team. Participants will be emailed a $50 gift card after completion of the focus group in recognition of their time. Purposive sampling will be by discipline i.e., all participants in each focus group will be from the same discipline to ensure representation from clinicians across institutions and with diversity in level of experience and primary discipline. All participants will be emailed a consent form (*online supplemental file 2*) and be asked to provide informed consent before participating in the focus group (*online supplemental file 3*).

### Data Collection

We developed a focus group guide (*online supplemental file 4*) based on the seven constructs included in the TFA (Table 1). We developed at least one question per domain, with prompts to probe domains for clarification or exploration. An ICU physician, registered nurses, registered respiratory therapists, and researchers reviewed the focus group guide for face validity. The focus group guide will be pilot tested with four groups of specialty-specific stakeholders (i.e., nurse practitioners, physicians, registered nurses, and registered respiratory therapists) from the *Venting Wisely* pilot implementation site (Foothills Medical Centre, Calgary AB) to refine wording and enhance clarity prior to conducting interviews.

**Table 1.** Theoretical framework of acceptability

| **Construct** | **Definition** |
| --- | --- |
| Affective attitude | How a clinician feels about the intervention |
| Perceived effectiveness | The extent to which the intervention is perceived as likely to achieve its purpose |
| Self-efficacy | The clinician's confidence that they can perform the behaviour(s) required to participate in the intervention |
| Burden | The perceived amount of effort that is required to participate in the intervention |
| Intervention coherence | The extent to which the clinician understands the intervention and how it works |
| Opportunity costs | The extent to which benefits, profits or values must be given up to engage in the intervention |
| Ethicality | The extent to which the intervention has good fit with a clinician's value system |

The Theoretical framework of acceptability proposes seven component constructs of acceptability.(18)

Focus groups will be moderated by a researcher (KK) or knowledge translation expert/registered nurse (AI) with experience in qualitative methods. A researcher will observe the focus groups and take notes to record details of participants surroundings, important features of participant responses, and themes to consider in the formal data analysis. Focus groups will be conducted remotely using Zoom. Duration of focus groups will be scheduled for 1.5 hours. Demographic data will be collected via an online survey (Qualtrics, Provo, UT) prior to the start of the focus group, including age, gender identity, ethnic, racial, or cultural self-identification, years of ICU experience, professional designation, and primary hospital site (*online supplemental file 5*). All focus groups will be audio-recorded, transcribed verbatim, verified, and deidentified. All focus group participants will be emailed a copy of the study report to review and comment upon as a form of member-checking.

### Sample Size

There are no a priori sample size considerations. We plan to conduct up to 17 focus groups at least two months post-implementation of the *Venting Wisely* pathway (see above *Participants and sampling frame*). Each focus group will consist of representatives from four prespecified ICUs from a single discipline. We will limit focus groups to eight clinicians for a total of up to 100 participants. We will conduct additional focus groups if needed to achieve theoretical saturation of themes (i.e., point when new data do not generate any new insights).

### Data analysis

De-identified transcripts will be imported into NVivo-12 (QSR International, Melbourne, Australia) for data management and analysis. Each participant group (i.e., nurse practitioner, physician, registered nurse, or registered respiratory therapist) will be analyzed independently to allow for the identification of discipline-specific themes. A coding template will be developed, with *a priori* themes based on the seven constructs of the TFA. Qualitative data will be collected and analyzed iteratively by two researchers (KK and AI). The researchers, working independently, will begin by reading the transcripts to gain familiarity with the content, followed by line-by-line inductive coding with constant comparison. The researchers will meet after reviewing every two to three transcripts to review emerging findings; differences will be resolved through discussion. The codes will then be mapped to the template of *a priori* themes, and additional themes emerging through the analysis will be added. Subthemes will be identified within and across themes. Once all transcripts are coded and mapped, the data will be organized to describe how participant experiences are aligned with and divergent from the TFA constructs (23-25). Quantitative demographic data will be summarized using descriptive statistics. The research team will meet regularly to review and discuss the findings. The multidisciplinary composition of the research team will ensure that the perspectives of all members of the ICU care team are reflected in the analysis and interpretation of data. Questions in the focus group guide may be adapted as focus groups are conducted and analyzed in order to further explore identified subthemes.

### Duration, Challenges, and Mitigation

We anticipate focus group guide refinement, recruitment of participants, focus group meetings and analysis will take up to 24 months. The largest risk will be challenges in recruitment. Our team will leverage our multi-disciplinary network of investigators and leaders to recruit clinicians as in previous studies completed successfully (26, 27).

### Knowledge Translation

We will use two types of knowledge translation throughout this study: integrated knowledge translation (iKT) and end of grant knowledge translation (28). Members of the ICU care team have been engaged throughout this study, from the development of the *Venting Wisely* pathway to development and refinement of the focus group guide. During data analysis, we will present our findings to *Venting Wisely* clinical advisors to evaluate and iteratively improve implementation of the *Venting Wisely* pathway at other ICUs and improve pathway adherence.

## ETHICS AND DISSEMINATION

### Ethics

This study was approved by the University of Calgary Conjoint Health Research Ethics Board (REB20-0646).

### Dissemination

We will compile a record of perceptions of the *Venting Wisely* pathway and how clinician involvement can be optimized and sustained. These will be included in a published report and inform future phases of this research program, including an exploration of the sustainability and (inter)national scalability of the Venting Wisely pathway. Study results will be shared with the 17 ICUs who participated in this study, submitted to a peer-reviewed journal for consideration of publication, and presented at a scientific conference. The results of the study will be disseminated to patients and the public at the completion of the trial.

### Conclusion

Exploring clinician experiences with the *Venting Wisely* pathway will contribute to a better understanding of the user’s experience of the *Venting Wisely* pathway. Study findings will be used to inform the refinement, implementation, and sustainment of the pathway to ensure its use is as intended, which in turn may improve outcomes of critically ill adults with HRF and ARDS. Study findings may also provide insights into how other complex interventions should and should not be implemented and adopted by multidisciplinary teams within an ICU setting.

**Figure 1.**
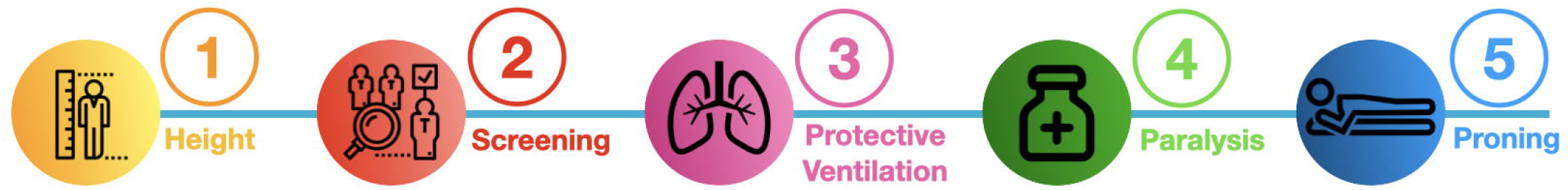
Five key steps of the Venting Wisely pathway.

## Data Availability

This is a protocol so there is no data associated with this manuscript. We have ethics approval to conduct this study.

## REFERENCES

1. Bellani G, Laffey JG, Pham T, Fan E, Brochard L, Esteban A, et al. Epidemiology, Patterns of Care, and Mortality for Patients With Acute Respiratory Distress Syndrome in Intensive Care Units in 50 Countries. JAMA. 2016;315(8):788–800.

2. Parhar KKS, Zjadewicz K, Soo A, Sutton A, Zjadewicz M, Doig L, et al. Epidemiology, Mechanical Power, and 3-Year Outcomes in Acute Respiratory Distress Syndrome Patients Using Standardized Screening. An Observational Cohort Study. Ann Am Thorac Soc. 2019;16(10):1263–72.

3. Bernard GR, Artigas A, Brigham KL, Carlet J, Falke K, Hudson L, et al. The American-European Consensus Conference on ARDS. Definitions, mechanisms, relevant outcomes, and clinical trial coordination. Am J Respir Crit Care Med. 1994;149(3 Pt 1):818–24.

4. Rubenfeld GD, Caldwell E, Peabody E, Weaver J, Martin DP, Neff M, et al. Incidence and outcomes of acute lung injury. N Engl J Med. 2005;353(16):1685–93.

5. Herridge MS, Tansey CM, Matté A, Tomlinson G, Diaz-Granados N, Cooper A, et al. Functional disability 5 years after acute respiratory distress syndrome. N Engl J Med. 2011;364(14):1293–304.

6. Herridge MS, Cheung AM, Tansey CM, Matte-Martyn A, Diaz-Granados N, Al-Saidi F, et al. One-Year Outcomes in Survivors of the Acute Respiratory Distress Syndrome. N Engl J Med. 2003;348(8):683–93.

7. Esteban A, Anzueto A, Frutos F, Alía I, Brochard L, Stewart TE, et al. Characteristics and Outcomes in Adult Patients Receiving Mechanical VentilationA 28-Day International Study. JAMA. 2002;287(3):345–55.

8. Luhr OR, Antonsen K, Karlsson M, Aardal S, Thorsteinsson A, Frostell CG, et al. Incidence and mortality after acute respiratory failure and acute respiratory distress syndrome in Sweden, Denmark, and Iceland. The ARF Study Group. Am J Respir Crit Care Med. 1999;159(6):1849–61.

9. Vincent J-L, Akça S, de Mendonça A, Haji-Michael P, Sprung C, Moreno R, et al. The Epidemiology of Acute Respiratory Failure in Critically Ill Patients. Chest. 2002;121(5):1602–9.

10. Cochi SE, Kempker JA, Annangi S, Kramer MR, Martin GS. Mortality Trends of Acute Respiratory Distress Syndrome in the United States from 1999 to 2013. Ann Am Thorac Soc. 2016;13(10):1742–51.

11. Walkey AJ, Goligher EC, Del Sorbo L, Hodgson CL, Adhikari NKJ, Wunsch H, et al. Low Tidal Volume versus Non-Volume-Limited Strategies for Patients with Acute Respiratory Distress Syndrome. A Systematic Review and Meta-Analysis. Ann Am Thorac Soc. 2017;14(Supplement_4):S271–s9.

12. Brower RG, Matthay MA, Morris A, Schoenfeld D, Thompson BT, Wheeler A. Ventilation with lower tidal volumes as compared with traditional tidal volumes for acute lung injury and the acute respiratory distress syndrome. N Engl J Med. 2000;342(18):1301–8.

13. Petrucci N, De Feo C. Lung protective ventilation strategy for the acute respiratory distress syndrome. Cochrane Database Syst Rev. 2013;2013(2):Cd003844.

14. Alhazzani W, Alshahrani M, Jaeschke R, Forel JM, Papazian L, Sevransky J, et al. Neuromuscular blocking agents in acute respiratory distress syndrome: a systematic review and meta-analysis of randomized controlled trials. Crit Care. 2013;17(2):R43.

15. Guérin C, Reignier J, Richard JC, Beuret P, Gacouin A, Boulain T, et al. Prone positioning in severe acute respiratory distress syndrome. N Engl J Med. 2013;368(23):2159–68.

16. Kohn LT, Corrigan J, Donaldson MS. To err is human : building a safer health system. Washington: National Academy Press; 2000.

17. Parhar KKS, Zjadewicz K, Knight GE, Soo A, Boyd JM, Zuege DJ, et al. Development and Content Validation of a Multidisciplinary Standardized Management Pathway for Hypoxemic Respiratory Failure and Acute Respiratory Distress Syndrome. Crit. 2021;3(5):e0428.

18. Sekhon M, Cartwright M, Francis JJ. Acceptability of healthcare interventions: an overview of reviews and development of a theoretical framework. BMC Health Serv Res. 2017;17(1):88.

19. Proctor E, Silmere H, Raghavan R, Hovmand P, Aarons G, Bunger A, et al. Outcomes for implementation research: conceptual distinctions, measurement challenges, and research agenda. Adm Policy Ment Health. 2011;38(2):65–76.

20. Parhar KKS. Identification and Treatment of Hypoxemic Respiratory Failure and ARDS With Protection, Paralysis, and Proning Pathway. 2023.

21. Parhar KKS, Soo A, Knight G, Fiest K, Niven DJ, Rubenfeld G, et al. Statistical analysis plan for the Identification and Treatment of Hypoxemic Respiratory Failure (HRF) and ARDS with Protection, Paralysis, and Proning: a type-1 hybrid stepped-wedge cluster randomized effectiveness-implementation study. medRxiv. 2023:2023.03.17.23287218.

22. Booth A, Hannes K, Harden A, Noyes J, Harris J, Tong A. COREQ (Consolidated Criteria for Reporting Qualitative Studies). Guidelines for Reporting Health Research: A User’s Manual2014. p. 214–26.

23. King N. Doing template analysis.. In: Symon G, Cassell C, editors. Qualitative organizational research : core methods and current challenges. London ;: SAGE; 2012. p. 426–50.

24. Brooks J, McCluskey S, Turley E, King N. The Utility of Template Analysis in Qualitative Psychology Research. Qualitative research in psychology. 2015;12(2):202–22.

25. Braun V, Clarke V. Using thematic analysis in psychology. Qual Res Psychol. 2006;3(2):77–101.

26. Stelfox HT, Niven DJ, Clement FM, Bagshaw SM, Cook DJ, McKenzie E, et al. Stakeholder Engagement to Identify Priorities for Improving the Quality and Value of Critical Care. PloS one. 2015;10(10):e0140141–e.

27. Bagshaw SM, Opgenorth D, Potestio M, Hastings SE, Hepp SL, Gilfoyle E, et al. Healthcare Provider Perceptions of Causes and Consequences of ICU Capacity Strain in a Large Publicly Funded Integrated Health Region: A Qualitative Study. Critical care medicine. 2017;45(4):e347–e56.

28. Gagliardi AR, Berta W, Kothari A, Boyko J, Urquhart R. Integrated knowledge translation (IKT) in health care: a scoping review. Implementation science : IS. 2016;11:38.

