## Supplemental files for "Acceptability of the Venting Wisely Pathway for use in Critically ill Adults with Hypoxemic Respiratory Failure and Acute Respiratory Distress Syndrome (ARDS): A qualitative study protocol"

**Contents**

Focus Group methods

1. Recruitment email ………………………………………………………………………………….. 1
2. Consent form …………………………………………………………………………………………. 2
3. Verbal consent form ………………………………………………………………………………. 6
4. Interview guide ……………………………………………………………………………………….. 8
5. Demographic survey ……………………………………………………………………………… 11

**1. Recruitment email**

Dear ICU clinician,

I am emailing to invite you to participate in a FOCUS GROUP to evaluate the acceptability of the TheraPPP (Venting Wisely pathway) that was recently implemented in your Intensive Care Unit. We need your expertise and opinions, to help us assess the acceptability of the TheraPPP (Venting Wisely pathway).

The focus groups would be conducted through your computer using Zoom videoconferencing platform
(https://zoom.us) through an institutional account and a link would be provided to you by our research team. The facilitator would ask you interviews on your perceptions of barriers and facilitators to conducting and implementing studies on new drugs that are related with long term outcomes in critically ill patients. The focus groups would be audio recorded for transcription purposes.

**The focus groups will take 1 to 1.5 hours to complete.** It would take place during a mutually agreed upon time with multiple critical care clinicians. To thank focus group participants for their time, we will be providing a $50 gift card.

Participation is voluntary. You will not be asked to provide any identifying data within the focus group. Should you choose to participate, the research team will do their best to make sure that your private information is kept confidential.

If you are interested in participating, more information will be provided. This study has been approved by the University of Calgary Conjoint Health Research Ethics Board (REB 20-0646).

We recognize how busy you are and greatly appreciate you considering participating in this focus group.

Sincerely,

Ken Parhar

Ken Parhar, MD, MSc, FRCPC
Clinical Associate Professor of Critical Care Medicine


### 2. Consent form

UNIVERSITY OF CALGARY

**TITLE:** Identification and Treatment of Hypoxemic Respiratory Failure (HRF) and ARDS with Protection, Paralysis, and Proning: TheraPPP (Focus Group – Venting Wisely pathway)

**INVESTIGATORS:** Dr. Ken Parhar (PI)

**SPONSOR:** Dr. Tom Stelfox (Department of Critical Care Medicine)
Canadian Institutes of Health Research (CIHR)

This information sheet is only part of the process of informed consent. It should give you the basic idea of what the research is about and what your participation will involve. If you would like more detail about something mentioned here, or information not included here, please ask. Take the time to read this carefully and to understand any accompanying information.

**BACKGROUND**

Acute hypoxemic respiratory failure (HRF) is common within the Intensive Care Unit (ICU). A significant proportion of these patients develop Acute Respiratory Distress Syndrome (ARDS), which is associated with a mortality of 40-60% in severe cases. Three interventions have been shown to improve the survival of patients with ARDS: Lung Protective Ventilation (LPV), neuromuscular blockade (paralysis), and prone positioning. The lack of a standardized approach outlining the management of ARDS patients has resulted in significant variability in the application of these interventions. Moreover, evidence suggests that the rational and algorithmic/pathway approach to the application of these interventions is associated with improved outcomes.

To bridge the gap between research and patient care, and deliver lifesaving therapies for HRF patients in a fair and rational way, the Department of Critical Care Medicine (DCCM) is implementing an evidence-based, stakeholder-informed care pathway (*Venting Wisely* pathway). The pathway was developed using a consensus process and relevant literature findings, and was validated by a broad group of stakeholders across Alberta. *Venting Wisely* standardizes the diagnosis and management of patients with HRF with the goal of reducing practice variation and improving adherence to evidence-informed therapy.

**WHAT IS THE PURPOSE OF THE STUDY?**

The purpose of this study is to evaluate effectiveness and acceptability of the *Venting Wisely* pathway. The specific purpose of this focus group is to evaluate clinician perceptions about the acceptability of the *Venting Wisely* pathway following implementation.

### WHAT WOULD I HAVE TO DO?

You are being asked to take part in a focus group to evaluate the acceptability of the *Venting Wisely* implementation. Because you are an ICU clinician who used the pathway, your feedback and input to pathway implementation is very important.

If you volunteer to participate in this study, the researcher will ask you to do the following:

- Complete a short demographic questionnaire in Qualtrics, an online survey platform, about you, your professional role, and hospital you work in.
- Join the focus group through your computer using the Zoom Videoconferencing platform (https://zoom.us) links provided to you by our research team. The focus group facilitator will provide you all the details you need to join the call prior to your focus group date.
- Participate in a discussion on the acceptability of the Venting Wisely pathway.
- Agree to have the study team record the audio of the focus group discussion. It is important to record the discussion so we can accurately document what it said.
- Once the interview is over, a member of the study team will send you a summary of the interview for you to review and provide feedback, should you want. The summary will be deidentified of participant personal information.

### HOW LONG WILL I BE IN THE STUDY?

Your participation in this study will take about 1.5 -2 hours.

### WHO ELSE WILL BE IN THE STUDY?

In each focus group, there will be up to to eight intensive care unit clinicians (Nurse Practitioners, Registered Respiratory Therapists, Registered Nurses, or physicians). Focus groups will include only one professional designation which means they will consist of only Nurse Practitioners, Registered Respiratory Therapists, or only Registered Nurses, or only physicians.

### WHAT ARE THE RISKS?

There are no foreseen risks to participating in this study. You will not be asked to provide any identifying data. All responses will be kept anonymous. Your confidentiality cannot be guaranteed as participants may not hold material confidential. Data will be presented in aggregate.

### WILL I BENEFIT IF I TAKE PART?

If you agree to participate in this study, there may or may not be a direct benefit to you. The information we gain from this process will be used to inform the full implementation of the pathway. The information you provide will also help with the design of other quality improvement initiatives. It is anticipated the results of this study will be shared with others in the following ways: medical journal articles, medical conferences, and summary report sheets for participants.

### DO I HAVE TO PARTICIPATE?

Participation in this study is voluntary. If you decide to take part, your consent to participate will be implied. You may decline to take part in this study, or at any time during the study you may decide to stop your participation without penalty. Once the audio recordings are transcribed it will be impossible to isolate individual participants which limits data withdrawal from the study at that point.

You will be advised in a timely manner of any new information that becomes available that may affect your willingness to remain in the study.

### WILL I BE PAID FOR PARTICIPATING, OR DO I HAVE TO PAY FOR ANYTHING?

To compensate your time, you will receive a $50 gift card for participating in this research study. After the focus group the facilitator will email participants a $50 "University of Calgary EverythingCard" redemption code. EverythingCard is Canada's most widely used gift card platform to simplify delivery of gift cards. The University of Calgary is using EverythingCard to allow researchers to purchase online codes for subject fees to be distributed to their research subjects as a token of appreciation for their time and effort in participating in a study. The EverythingCard platform allows participants to redeem their codes online and select their own gift card(s) from a variety of retailers.

### WILL MY RECORDS BE KEPT PRIVATE?

Your privacy is important to us. The information collected will be stored and maintained confidentially and destroyed as required by law. Your name and personal information will not be made available to anyone who is not involved in this study unless disclosure is required by law. The results of this study will be published in a medical literature, but your personal information will not be revealed.

Your demographic data will be collected in Qualtrics an online survey platform with servers located in Toronto, Ontario, Canada. All data are encrypted and stored directly on its servers. Researcher access to the survey data is password-protected and the transmission is encrypted. IP tracking will be off. Survey responses cannot be linked to your computer. All information will be stored in a secured area (i.e. locked filing cabinet and/or password protected computer).

A third-party transcription service, Rev.com, will be used to transcribe the focus group interviews. Rev.com is an online transcription service that follows best practices handling personally identifiable information with guidance from the published General Data Protection Regulation. Information regarding their privacy policy can be found at https://www.rev.com/about/privacy.

### AGREEMENT TO PARTICIPATE

Your participation in the focus group will be interpreted as explicit oral consent of your agreement to participate. In no way does this waive your legal rights nor release the investigators, or involved institutions from their legal and professional responsibilities. You are free to withdraw from the study at any time.

If you have further questions concerning matters related to this research, please contact:

Gwen Knight, Research Associate (403) 944-0735
Or
Dr. Ken Parhar (403) 944-0735

If you have any questions concerning your rights as a possible participant in this research, please contact the Chair of the Conjoint Health Research Ethics Board, Research Services, University of Calgary, 403-220-7990.

The University of Calgary Conjoint Health Research Ethics Board has approved this research study.

**3. Oral consent form**

**UNIVERSITY OF CALGARY**
**ORAL CONSENT TO PARTICIPATE IN RESEARCH**

Thank you for agreeing to participate in today's focus group.

My name is [insert name] and I am a [role] at the University of Calgary working on this study.

The purpose of this study is to evaluate effectiveness and acceptability of the *Venting Wisely* pathway. The specific purpose of this focus group is to evaluate clinician perceptions about the acceptability of the *Venting Wisely* pathway following implementation.

You are being asked to take part in a focus group to evaluate the acceptability of the *Venting Wisely* pathway implementation. Because you are an ICU clinician who used the pathway, your feedback and input to pathway implementation is very important.

If you volunteer to participate in this study, the researcher will ask you to do the following:

- Complete a short demographic questionnaire in Qualtrics, an online survey platform, about you, your professional role, and hospital you work in.
- Join the focus group through your computer using the Zoom Videoconferencing platform (https://zoom.us) links provided to you by our research team. The focus group facilitator will have provided you with the details you need to join the call prior to your focus group date.
- Participate in a discussion on the acceptability of the Venting Wisely pathway.
- Agree to have the study team record the audio of the focus group discussion. It is important to that we record the discussion so we can accurately document what it said.
- Once the interview is over, a member of the study team will send you a summary of the interview for you to review and provide feedback, should you want. The summary will be deidentified of participant personal information.

Your participation in this study will take about 1.5 -2 hours.

Your privacy is important to us. Your name and personal information will not be revealed to anyone who is not involved in this study. Confidentiality is kept unless disclosure is required by law; however, your confidentiality cannot be guaranteed as participants may not hold material confidential. In addition to four to eight single profession participants, only the focus group facilitator and notetaker will be present. Audio recordings will be transcribed to preserve the details of our discussion. All potentially identifying information (ex. names, locations of hospitals or cities) will be removed to de-identify the data. The results of this study will be published in the medical literature. Your personal information will not be revealed. The information collected will be stored and maintained confidentially as required by law.

Your demographic data will be collected in Qualtrics an online survey platform with servers located in Toronto, Ontario, Canada. All data are encrypted and stored directly on its servers. Researcher access to the survey data is password-protected and the transmission is encrypted. IP tracking will be off. Survey responses cannot be linked to your computer. All information will be stored in a secured area (i.e. locked filing cabinet and/or password protected computer).

A third-party transcription service, Rev.com, will be used to transcribe the focus group interviews. Rev.com is an online transcription service that follows best practices handling personally identifiable information with guidance from the published General Data Protection Regulation. Information regarding their privacy policy can be found at https://www.rev.com/about/privacy.

You may contact Dr. Ken Parhar at (403)-944-2471 with any questions or concerns about the research or your participation in this study.

If you have any questions concerning your rights as a participant in this research, please contact the Chair, Conjoint Health Research Ethics Board, University of Calgary at 403-220-7990.

Do you have any questions or would like any additional details? [Answer questions.]

Taking part in this study is your choice. You can choose whether or not you want to participate. Whatever decision you make, there will be no penalty to you. You have a right to have all of your questions answered before deciding whether to take part. If you decide to take part, you may leave the study at any time.

In no way does your agreement to take part this study waive your legal rights nor release the investigators or involved institutions from their legal and professional responsibilities.

Do you agree to participate in this study?

[If yes, begin the study.]

[If no, thank the participant for his/her time.]

### 4. Interview guide

#### Briefing (5 minutes)

1. Welcome and thank you for agreeing to take part in this focus group discussion about the Venting Wisely pathway.
2. [Introduce self]
3. As described in our email, we are interested in hearing about your perceptions of the Venting Wisely pathway as well as feedback on what can be done to improve and sustain its implementation.
4. We emailed you a copy of the informed consent form. The consent form is part of the process of informed consent. It should give you a basic idea of what the research is about and what your participation will involve. Did everyone receive the consent form and have a chance to read through it Good. Because it is important that you understand your rights as a participant, I just want to review the main elements in the consent form:

[Interviewer will read the REB approved Oral Consent Script]

5. All information I collect is confidential. I hope this encourages you to speak freely. We would like the discussion to be informal, so there's no need to wait for us to call on you to respond. In fact, we encourage you to respond directly to the comments other people make. If you don't understand a question, please let us know. We are here to ask questions, listen, and make sure everyone has a chance to share.
6. Does anyone object to me recording our conversation? The recording will be typed out, but everything you say will be anonymous.

[Press record]

**Ground rules**

- The most important rule is that only one person speaks at a time.
- There are no right or wrong answers
- You do not have to speak in any particular order
- You do not have to agree with the views of other people in the group
- Does anyone have any questions? (answers).
- Ok let's begin:

You have been asked to participate in this study because you work in an ICU where the Venting Wisely pathway was implemented. Briefly, this was an evidence-based, stakeholder-informed pathway for the diagnosis and management of hypoxemic respiratory failure. This pathway included six steps: 1) Measure: where all mechanically ventilated patients had their height and predicted body weight measured and recorded; 2) Screen: Where patients were screened for the presence of HRF using PF ratios. 3) Manage: Lung Protective Ventilation was initiated for patients with HRF; 4) Monitor: plateau pressure & driving pressure, optimal PEEP study; 5) Paralysis: if the patient develops worsening HRF and does not meet LPV goals, therapy was escalated using a neuromuscular blockade. Patients with worsening PF ratio despite steps 1-4 will be considered for prone positioning followed by proning, followed by ECLS.

**Warm up**

| I'd like everyone to introduce themselves. Please tell us your name and a sentence or two about the background that brings you here | | |
|---|---|---|
| **Question** | **Probe** | **Notes** |

| | |
|---|---|
| 1. I'm going to give you a minute to think about your experience as a clinician using the Venting Wisely (VW) pathway: How do you feel about the Venting Wisely pathway?<br>*Affective Attitude: How an individual feels about an intervention* | What did you like about it?<br>(Ask about implementation or the pathway itself)<br>What did you dislike about it?<br>(Ask about implementation or the pathway itself) |
| 2. How do you think using the Venting Wisely pathway will impact the use of evidence-based care and improve patient outcomes?<br>*Perceived effectiveness: The extent to which the intervention is perceived as likely to achieve its purpose* | If yes, what do you think some of the benefits of the VW are?<br><br>If not, is this due to a problem with implementation of the pathway on your unit? Or with the pathway itself? (maybe ask more neutral as this is leading: could just ask why and ask probing questions to understand) |
| 3. How did the Venting Wisely pathway change your confidence in caring for HRF / ARDS patients?<br>*Self-efficacy: The participant's confidence that they can perform the behaviour(s) required to participant in the intervention* | What's your confidence level that you can perform all of the Venting Wisely pathway therapies associated with your discipline?<br>What kind of support was helpful in gaining confidence?<br>• Implementation support (e.g. education at Grand Rounds)?<br>• Experience with the pathway post implementation?<br>Is there something that **could have** supported you in gaining confidence?<br>• During implementation?<br>Within the pathway itself? |
| 4. How much time or effort was it to use the Venting Wisely pathway?<br>*Burden: The perceived amount of effort that is required to participate in the intervention* | If a lot, ask about implementation or the use of the pathway itself.<br>If not much, did it make your day more efficient? |
| 5. What is your understanding of the goal of the Venting Wisely pathway and how it | What was (or would be) most helpful ( for you to understand the goals of VW and how to |

| | |
|---|---|
| works?<br>*Intervention coherence: The extent to which the participant understands the intervention and how it works* | perform the pathway? |
| 6. What was it like to balance the Venting Wisely pathway with all the other daily tasks for your patients?<br>*Opportunity costs: The extent to which benefits, profits, or values must be given up to engage in the intervention* | Were there patient care tasks you felt you had to give up, or do more quickly, to incorporate VW pathway therapies?<br><br>If yes, what part of the pathway took too much time and what did you end up giving up (pathway or other)? |
| 7. Do you think the Venting Wisely pathway is in the best interest of patient? What about the best interests of the provider?<br>*Ethicality: The extent to which the intervention has good fit with an individual's value system* | Which parts are in the best interests of patient or provider?<br>Which parts are NOT in the best interests of patient or provider? |
| 8. Our aim is that VW becomes part of daily clinical practice. What could help this sustained once the project team is no longer available? | Which elements of the path will be most difficult to sustain? |
| Optional, time permitting: What did your unit do really well in adopting Venting Wisely? | Why do you think you were so successful? |
| Is there anything else you would like to share with us? | If no one answers, go around and ask what is one thing you would like to tell the study team about the Venting Wisely pathway? |
| Do you have any questions for us? | |

**Summary** (depending on time)

Do you have any questions for us?

Thank you for sharing your time and personal experience!

5. **Demographic survey**

**Post focus group – Demographic Survey**

Thank you for participating in the focus groups for the “Identification and Treatment of Hypoxemic Respiratory Failure (HRF) and ARDS with Protection, Paralysis, and Proning: TheraPPP” study. We want to record further demographic information about our participants. We appreciate your consideration of this request.

The questions will take a couple of minutes to complete. Your responses will remain confidential and stored in password-protected file on secure server at the University of Calgary. Only members of our research team will have access to the information.

If you are not comfortable answering any of the below questions you are welcome to skip any or all of those you do not wish to answer. Thank you for your time and continued participation.

### <u>Demographics</u>

1.. What is your gender identity?

Man

Woman

Non-binary

Transgender

Two-Spirit

Prefer not to answer

Prefer to self-identify

2. What is your age?

3. Which ethnic, racial, or cultural group do you most closely self-identify with?

*Note that the examples provided are non-exhaustive and are meant to be a guide to help you respond to the question. (Please select all that apply)*

Arab/West Asian (e.g., Egyptian, Iranian, Lebanese)

Black

Caucasian

Chinese

Indigenous Peoples

Japanese

Korean

Latin American (e.g., Argentinian, Chilean, Salvadorean)

South Asian (e.g., Indian, Pakistani, Sri Lankan)

Southeast Asian (e.g., Malaysian, Filipino, Vietnamese)

Prefer to describe

Prefer not to answer

4. How many years have you worked in your current role?

< 1 year

1-2 years

3-5 years
6-10 years
11-20 years
>20 years

5. [If Clinician] How big is the population your hospital serves?

<50,000

50,000-100,000

100,000 – 500,000

500,00 – 1,000,000

1,000,000 – 2,500,000

2,500,000 – 5,000,000

5,000,000 – 10,000,000

Other (please specify)

6. How many beds in total does your hospital have?

<250

250-500

500-1000

>1000

Other (please specify)

7. How many beds in total does your ICU have?

8. What is your professional designation?

9. What is your primary role in the intensive care unit?:

Intensivist

Base specialty

Critical Care Fellow

Base specialty

Resident

Base specialty

Registered Nurse

Registered Respiratory therapist

Other clinician

10. The hospital I work at:

11. How many years of experience do you have working in an intensive care unit/ critical care medicine?

<1 year
1-2 years
3-5 years
6-10 years
11-20 years
>20 years
